# Assessing the Risk of Depression Among U.S. Adults with Resolved Thyroid Dysfunction--data analysis from the National Health and Nutrition Examination Survey (2013–2018)

**DOI:** 10.1101/2024.05.07.24306885

**Authors:** Denghuang Zhan, Pinyu Chen, Ehsan Karim

**Affiliations:** School of Population and Public Health, the University of British Columbia, Vancouver, BC, Canada; School of Medicine, Royal College of Surgeons Ireland, Dublin, Ireland

## Abstract

**Background:** Depression is a highly-prevalent disease among US adults. The positive association between thyroid dysfunction and depression has been identified in many studies, but whether the thyroid dysfunction history (have recovered) is still associated with depression, and whether this association is modified by gender is still unknown. This finding can be important for depression preventions.

**Methods:** We applied design-adjusted multivariable logistic regression to examine the adjusted association between thyroid dysfunction history and depression by using data from the National Health and Nutrition Examination Survey from 2013 to 2018.

**Results:** Of 11975 respondents with complete responses included in this study, 8.4% (n = 1007) of the respondents have depression. The design-adjusted analysis shows no significant association between the thyroid dysfunction history and depression but shows gender as a significant effect modifier of the association. The association is 3.31(odds ratio, 95% CI: [1.38,7.93]) for males and 1.15 (odds ratio, 95% CI: [0.72, 1.84]) for females.

**Conclusion:** People who had thyroid dysfunction but recovered will still have a higher risk of getting depression, and it differs in genders. More suggestions and actions are needed for those who recovered from the thyroid dysfunction in order to prevent depression, especially for the males.

## Introduction

### Background and Rationale

Depression is one of the leading cause of disability and affecting more than 350 million people worldwide (1, 2). In US, the study showed that one in five individuals would experience depressive symptoms at least once in their lifetime (3). A recent study even shows that depression prevalence has increased significantly from 8.7% in 2017–2018 to 14.4% in April 2020 (4). Thyroid dysfunction is one of the most common endocrine disorders during clinical practice and the prevalence of thyroid dysfunction varies by age, sex, race/ethnicity, and dietary iodine intake (5, 6). Thyroid dysfunction has important impacts on health outcomes including cardiovascular arrhythmia, metabolism, bone health, as well as mental health (7–10).

The association between thyroid function and psychiatric disorders has long been recognized, but it is only extensively studied in the most recent thirty years (11). In the late 1960s, the link between thyroid hormones and depression was identified in a clinical cohort where the analysis showed that the efficiency of antidepressant treatment for the depression patients was higher after delivery of triiodothyronine (12), and this finding was also confirmed by several clinical trials (13–15). The link between the thyroid dysfunction and the depression were then better understood after more studies have realized the neurobiological effects of the hypothalamic-pituitary-thyroid (HPT) axis in mood modulation, and the role of thyroid function in the pathophysiology (16–19). These studies show that the activation of the HPT axis involves hypothalamic release of thyrotropin-releasing hormone (TRH), which will further stimulate the release of thyroid-stimulating hormone (TSH) from the pituitary gland and, finally, activates the release of the thyroid hormones. With this relationship, thyroid dysfunction has been the focus of studies evaluating the effects of thyroid dysfunction on mood disorders and the association between TSH and depression has been consistently demonstrated in the literature (20–22). Now it is generally believed that the basic mechanism of thyroid dysfunction affecting mental status is through excess or insufficient released thyroid hormones and adequate thyroid treatments would alleviate depression symptoms caused by thyroid dysfunction (12, 23). However, to our knowledge, there are no studies showing if the thyroid dysfunction history has effects on the depression, i.e., if the patients still at higher risks of having depression even after they have recovered from thyroid dysfunction.

### Study aim

This cross-sectional study aims to assess whether the patients who had thyroid dysfunction history are still at a higher risk of depression than those who never had the thyroid dysfunction before among US adults. The hypothesis is that there is a significant association between the thyroid dysfunction history and depression. Also, one recent study shows that the gender may play a substantial role in the relationship between TSH and depressive symptoms (24), our second aim is thus to assess if gender is an effect modifier of this association. The hypothesis is the association between the thyroid dysfunction history and the depression differs in male and female groups. We will try to identity the two aims by applying multivariable logistic regression with careful considerations of the covariates adjustments, using National Health and Nutrition Examination Survey (NHANES) as data source.

The findings from this study may help the health organizations with their policies/suggestions makings to prevent depression, with thyroid dysfunction history taken into considerations.

## Method

### Data source, Design and Study population

The National Health and Nutrition Examination Survey (NHANES) includes a series of cross-sectional nationally representative health examination surveys. In each survey cycle, a nationally representative sample of the US civilian noninstitutionalized population is selected using a complex, stratified, multistage probability cluster sampling design (25, 26). A detailed description of the survey design, sample frame, and interviewing procedures can be obtained from the NHANES website (26). Ethics is covered by the University of British Columbia’s Policy 89, item 7.10.3 on studies involving human participants and Article 2.2 of the Tri-Council Policy Statement on Ethical Conduct for Research Involving Humans (TCPS 2) (27).

In this cross-sectional study, we used the data from the cycle 2013 to 2018. A total of 29400 individuals were included in the survey during the time period.

### Analytic sample and Study variables

For all the variables considered in this study, the responses such as ‘Don’t Know,’ ‘Refusal’, ‘Missing’ or ‘Not Stated’ were all considered as missing data. Only individuals aged 20 and above, not having thyroid dysfunction at the time when taking the survey in the cycle 2013 to 2018, and not having missing data in the exposure/outcome were included. Figure 1 summarizes the exclusion/inclusion restrictions for creating the analytic sample and demonstrates the corresponding sample sizes at each step.

**Figure 1:**
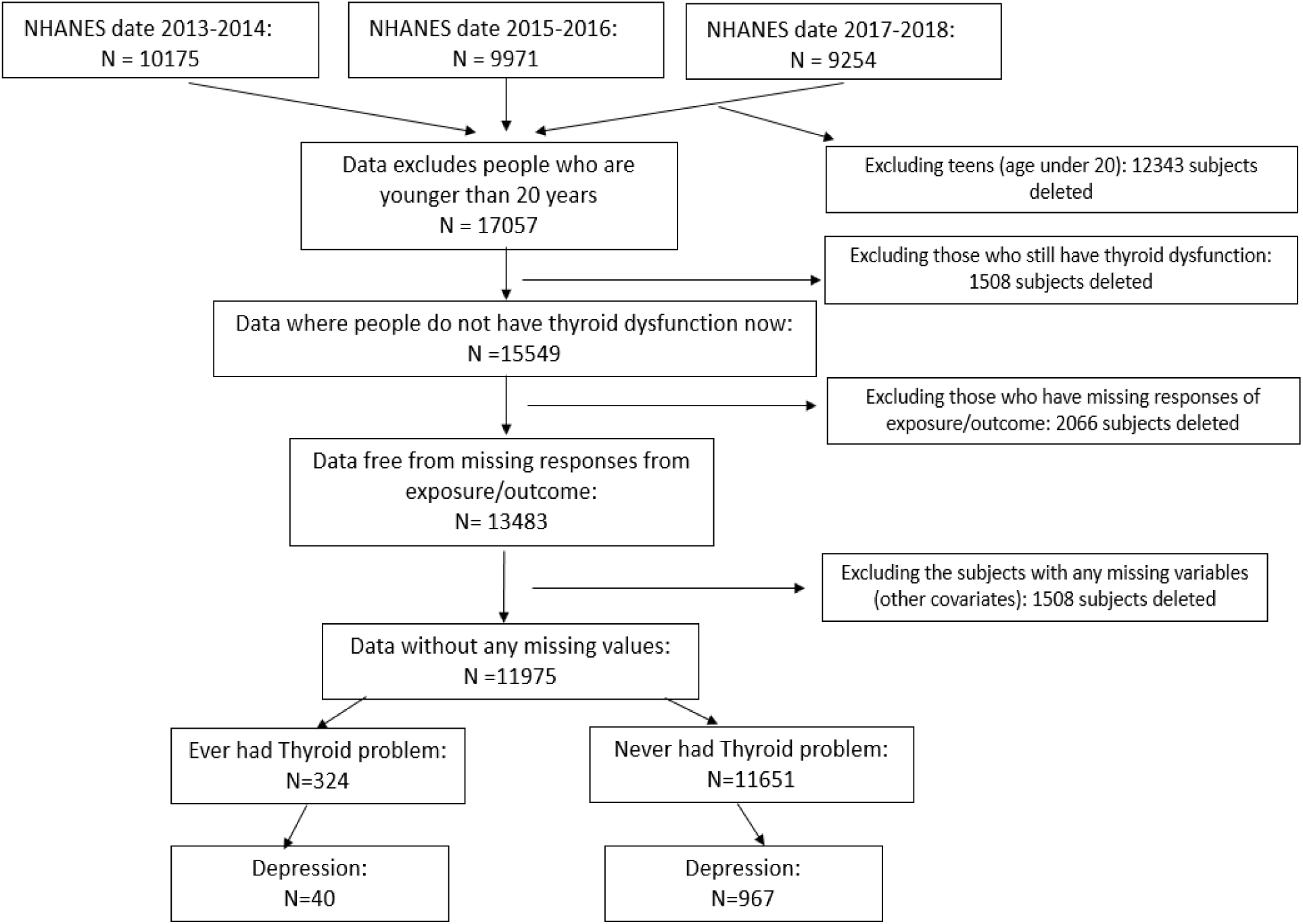
Flow chart of the analytic sample for the association between the thyroid dysfunction history and depression in US adults from the National Health and Nutrition Examination Survey (NHANES), United States, 2013–2018 for primary, secondary and sensitivity analysis. Specifically, the complete case dataset contains n=11975 unweighted samples, and the dataset free from missing responses in exposure/outcome contains n=13483 unweighted samples.

Specifically, the binary outcome variable for depression (yes/no) over the past 2 weeks was obtained by the 9-item Patient Health Questionnaire (28) (also known as PHQ-9). The total score of 9 was used as the cut-off (29), i.e., a total score larger than 9 was considered as having depression.

The exposure variable was the thyroid dysfunction history (yes/no), and it was obtained directly form a survey question “Ever told you had thyroid problem?”.

Based on a *prior* knowledge, we built the potential directed acyclic graph (DAG, shown in Figure 2), with the following potential confounders identified based on literatures (6, 18, 24, 30, 31): age, gender, income level, marriage, body mass index (BMI), hypertension, physical activity, race, health insurance, diabetes, smoking status and education level. These variables were renamed and re-leveled for the study from the NHANES.

**Figure 2:**
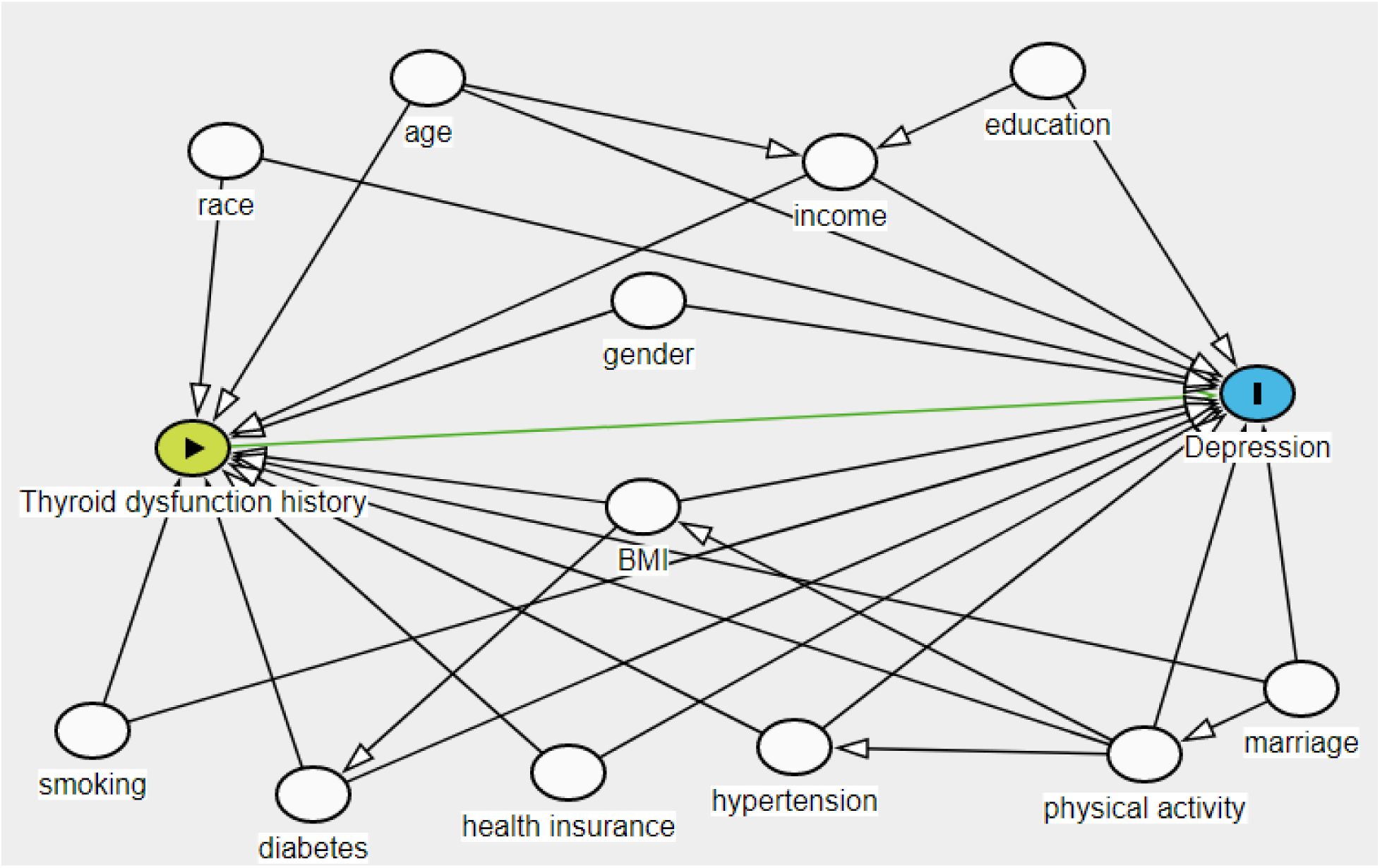
The directed acyclic graph for assessing the effect of thyroid dysfunction history on depression from the National Health and Nutrition Examination Survey, 2013-2018. All following covariates in the graph were renamed and re-leveled for the study: age (categorized as 20-39 years, 40-59 years, 60-79 years and 80 years and above), gender (males/ females), annual household income level (less than $20,000, $20,000 to $54,999, $55,000 and more), education level (low for “Less than 9th grade” or “9-11th grade” , medium for “High school graduate/GED or equi” or “Some college or AA degree”, high for “College graduate or above”), marriage ( married, never married and previously married ), self-reported smoking status (never smoked, former smoker and current smoker), body mass index (BMI) (underweight if smaller than 18.50, normal weight if between 18.5 and 24.99, overweight if 25.00 and above), hypertension (yes/no), physical activity level (inactive/active), race (white, black, Asian, Hispanic and other), health insurance (yes/no) and diabetes (yes/no).

### Statistical analysis

#### Descriptive statistics

For the complete case sample, the characteristics of the sample were demonstrated as unweighted numbers and as a weighted sample percentage, representing the population-level percentage. The sample was stratified by the outcome depression, and Rao–Scott χ2 test (32) was used to calculate the between-group differences in terms of the depression.

#### Primary analysis

The primary analysis was focused on complete case dataset. The design-adjusted multivariable logistic regression was used to evaluate the association between the thyroid dysfunction history and depression at the US population level. The associations between the outcome and covariates were reported as odds ratio (OR) together with the confidence interval (CI), and the standard errors of the point estimates were estimated by Taylor series linearization.

Based on the DAG, the minimal sufficient adjustment set including age, gender, income level, marriage, body mass index (BMI), hypertension, physical activity, race, health insurance, diabetes and smoking status were then fixed in the regression model. The variable ‘education level’, which is a confounder based on conventional definition (33) but not in the minimal sufficient adjustment set, was selected through backward elimination based on the Akaike Information Criterion (AIC). The variable selection procedure was also conducted based on the consideration of collinearity (VIF larger than 3), but this variable selection procedure was only conducted on the variable ‘education level’.

To identify the potential interactions between gender and thyroid dysfunction history, Analysis of Variance (ANOVA) test was used to check if the interaction term is significant among the two nested models. The goodness-of-fit for the final determined model was evaluated by Archer and Lemeshow test (34) and the model discrimination was assessed by weighted Receiver Operating Characteristic (ROC) curve.

#### Secondary analysis

The secondary analysis was focused on the imputed dataset. Specifically, we used multiple imputation by chained equations to provide missing data on the variables with missing values especially the ‘income’ (10% missing rate). Under the assumption of missing at random, predictors to be put in the imputation models were selected by a minimum correlation of 5 % among all the variables shown in DAG (35). Missing data were imputed in five datasets with predictive mean matching, the number of iterations was set to be 10. A survey-weighted logistic regression model, which was determined by the primary analysis, was fit to each imputed dataset to estimate the effect of thyroid dysfunction history on depression. The results were pooled by Rubin’s rules (35).

#### Sensitivity analysis

The sensitivity of the primary and secondary analysis was evaluated by propensity score matching analysis (36). Following the Zanutto’s (37) suggestion, we did not include any survey features but put all known confounding variables in the DAG into the propensity score logistical regression model. The subjects were then matched by propensity score without replacement with a ratio 1: 4, using the nearest neighborhood approach with caliper bound of 0.2 times the standard deviation of the propensity score. The covariate balance in the matched sample was measured by standardized mean difference (SMD) (38). unbalanced covariates after matching (SMD > 0.2) were adjusted for in the outcome model. The sensitivity analysis was performed in both the complete case dataset and the imputed datasets.

All statistical analyses were performed using R-4.1.1(39). For all the tests, a p-value of less than 0.05 was considered to indicate statistical significance.

## Results

### Descriptive statistics

The complete case sample includes n =11975 subjects, with 2.7% (n = 324) of the respondents having thyroid dysfunction history and 8.4% (n = 1007) of the respondents having depression. Table 1 summarizes the descriptive statistics for all covariates stratified by the outcome depression. The crude association between the thyroid dysfunction and depression is found to be 1.71 (odds ratio, 95% CI [1.07;2.75]). In Table 1, the survey-sample number is unweighted, while the sample percentage is weighted to represent the US population percentage.

**Table 1.**
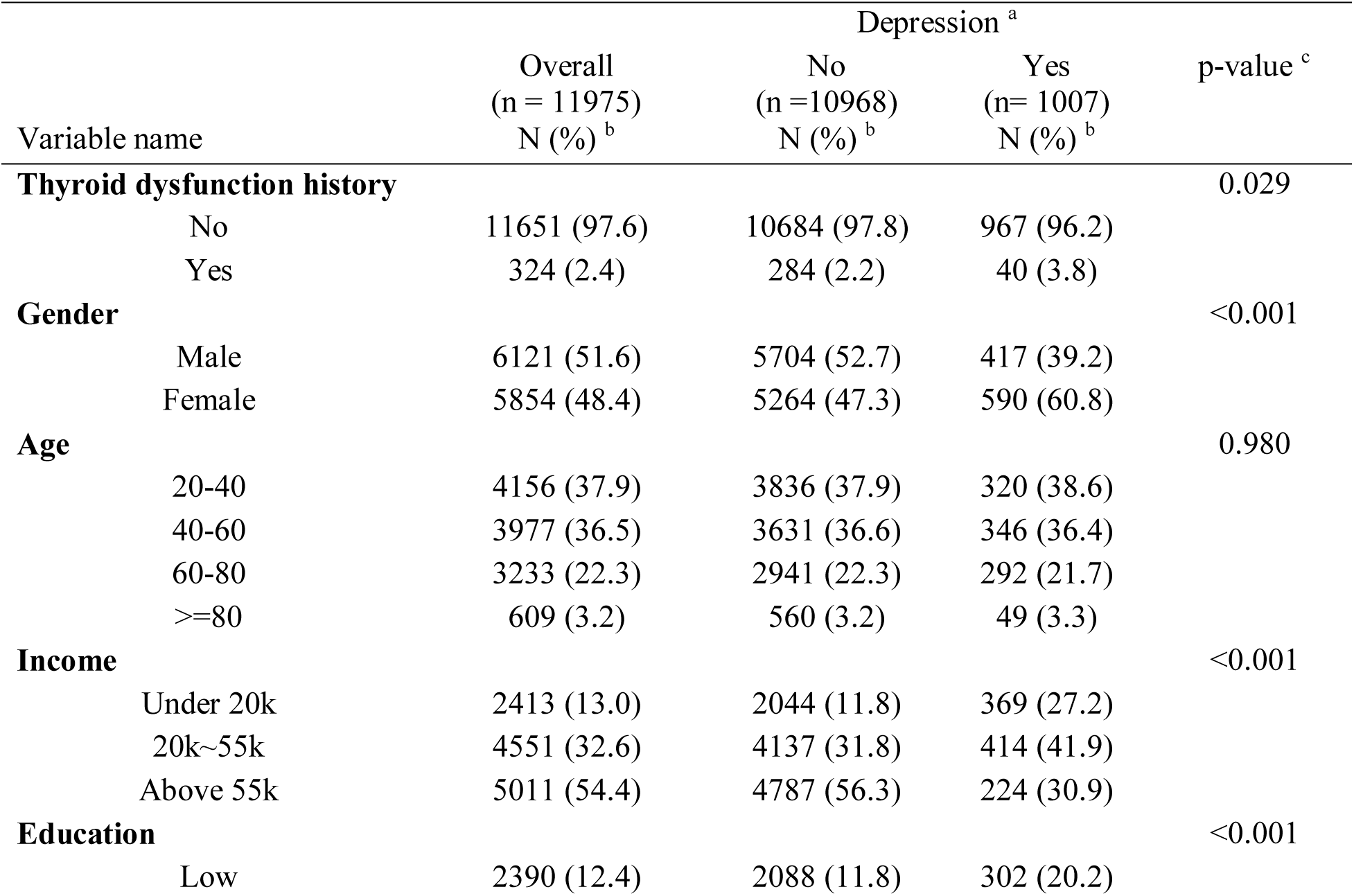
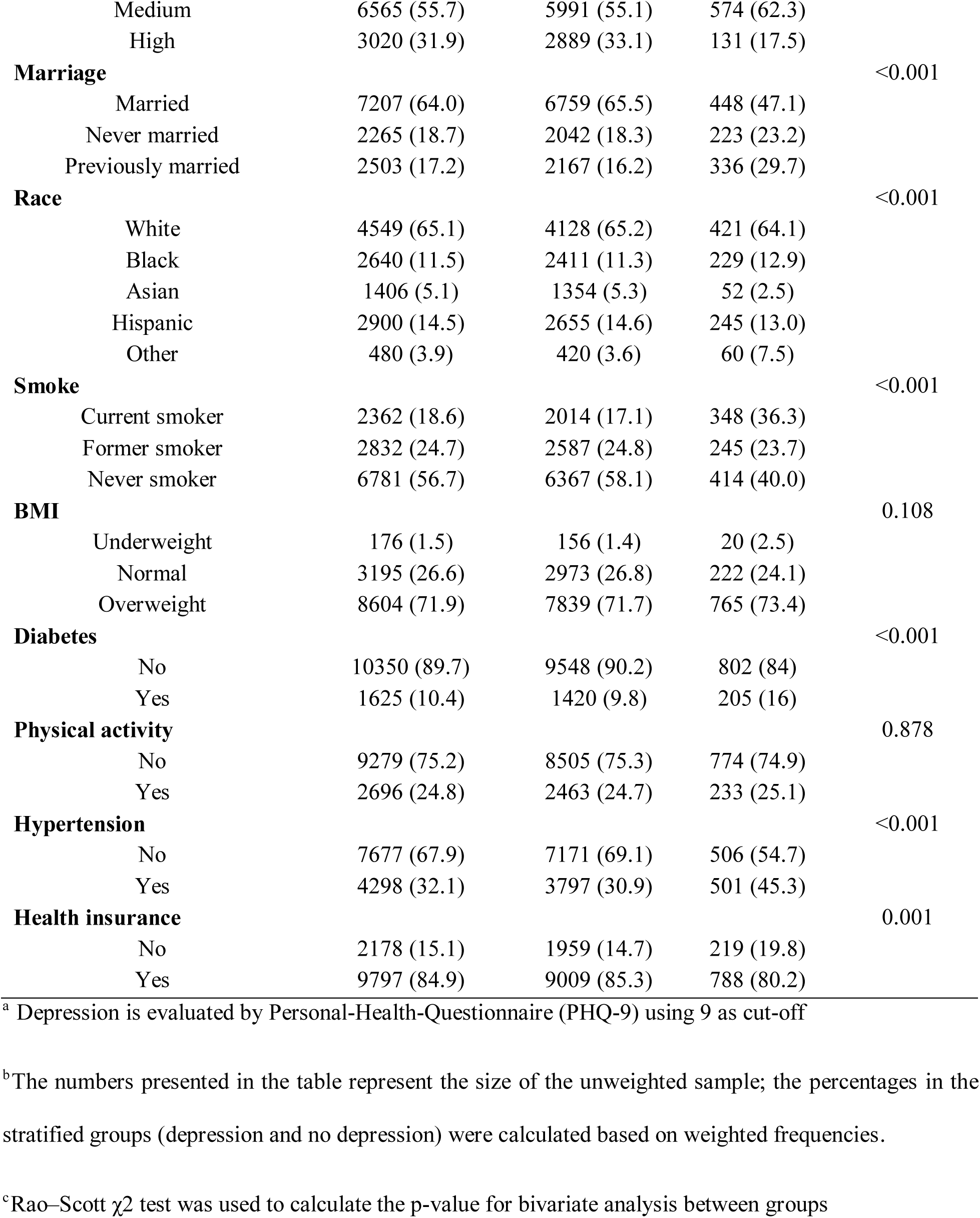
Univariate and bivariate statistics of study participants stratified by the outcome depression; National Health and Nutrition Examination Survey (NHANES), United States, 2013–2018.

### Primary analysis

The design-adjusted multivariable logistical regression model was first built by including all potential confounders as described in the DAG. The AIC backward elimination procedure shows that including the variable education in the model increases the performance of the model fit, the variable education was thus kept in the model. Also, no collinearity issue was detected for any covariates with a VIF cut-off point of 3. According to the model outputs, the odds of having depression was 1.44 (95% CI: [0.90;2.32]) times higher among those who had thyroid dysfunction history than those did have. The ANOVA test shows that the models with and without the thyroid dysfunction history-gender interaction term differ significantly (p-value = 0.039), indicating the gender is an effect modifier in this association. The main outputs from this final determined regression model for males and females are summarized in Table 2. From Table 2, the males who had the thyroid dysfunction history have 3.31 (95% CI:[1.38,7.93]) times the odds of getting depression than those never had the thyroid dysfunction while this odds ratio is 1.15 (95% CI: [0.72, 1.84]) for the females.

**Table 2:**
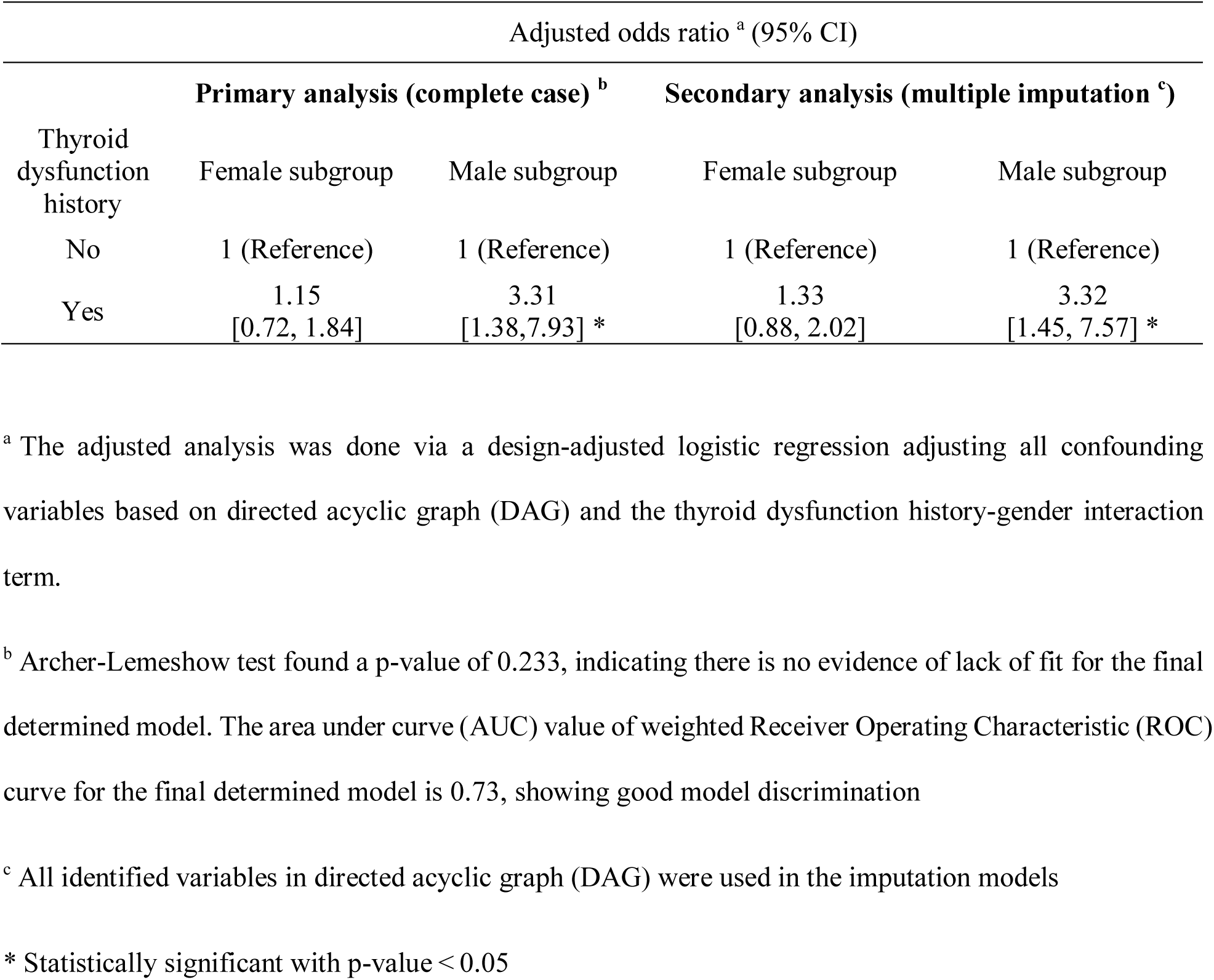
The associations between thyroid dysfunction history and depression in males and females, from primary analysis (complete case) and secondary analysis (multiple imputation); National Health and Nutrition Examination Survey (NHANES), United States, 2013–2018

### Secondary analysis

The association between thyroid dysfunction history and depression was also investigated by multiple imputation of missing values in some variables. The minimum correlation matrix selected all available variables for the imputation models. A total 1508 observations were added by multiple imputation. The final determined logistical regression model (with thyroid dysfunction history-gender interaction term) from primary analysis was used for the secondary analysis as the primary analysis shows the gender is an effect modifier. Combined estimates from the five imputed datasets show that those with thyroid dysfunction history have 3.32 (95%CI: [1.45–7.57]) times the odds of getting depression than those never had thyroid dysfunction history in males and the odds ratio is 1.33 (95%CI: [0.88, 2.02]), in females as shown in Table 2.

### Sensitivity analysis

The sensitivity of the primary and secondary analysis was tested by propensity score matching. This process was done separately in male and female groups because the primary analysis identified gender as an effect modifier. The SMD of the covariates in the matched sample all met the < 0.2 cut-off; therefore, none of them were adjusted for in the outcome model. As shown in Table 3, the propensity score analysis for the complete case dataset shows that odds ratio is 3.11(95% CI:[1.21 ,9.59]) in males and 1.21 (95% CI:[0.78, 2.35]) in females, while for imputed datasets, the odds ratio is 3.12 (95% CI:[1.26 ,9.19]) in males and 1.25 (95% CI:[0.84, 2.25]) in females.

**Table 3:**
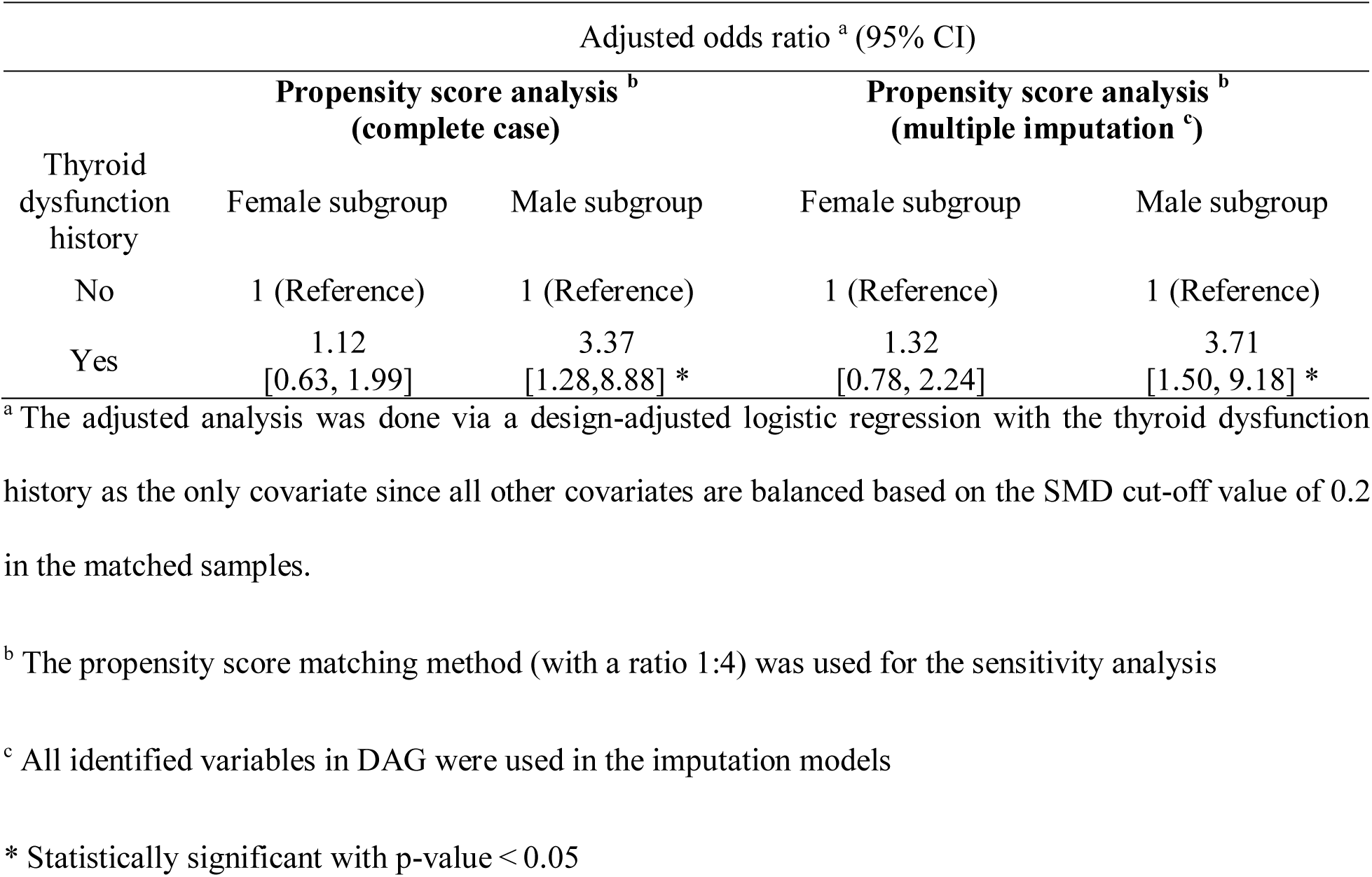
The associations between thyroid dysfunction history and depression in males and females, from sensitivity analysis using complete case dataset and imputed datasets; National Health and Nutrition Examination Survey (NHANES), United States, 2013–2018

We do not interpret effect estimates for variables other than the primary exposure, as the interpretation of other covariates may require specification of a separate causal framework to account for additional confounding (40).

## Discussion

### Interpretation

The primary analysis showed that the odds of having depression was 1.44 (95% CI: [0.90;2.32]) times higher among those who had thyroid dysfunction history than those did have, after adjusting for the confounders: age, race, income, education, marriage, smoking status, BMI, hypertension, physical activity, health insurance and diabetes, but without taking the thyroid dysfunction history-gender interaction term into consideration. However, gender was found to be a significant effect modifier in estimating the association between the thyroid dysfunction history and depression in the population of the United States, indicating that the results from the model without interaction term were not meaningful anymore. Our primary analysis thus further showed that, among American males, people with thyroid dysfunction history had 3.31 times the odds of getting depression than those who never had thyroid dysfunction before (OR= 3.31, 95% CI: [1.38,7.93]); while among females, this odds ratio was found only to be 1.15 (OR =1.15, 95% CI: [0.72, 1.84]) and not significant. The secondary analysis (multiple imputation) and sensitivity analysis (propensity score matching) in general agreed with the primary results, indicating that the thyroid dysfunction history would still impact the development of depression, and the impact for males was much stronger than females.

To the best of our knowledge, this is the first study investigating the associations between the thyroid dysfunction history and the depression among US people and examining if this association is modified by gender. Due to this reason, it is hard to find the articles that are examining the same questions in this context. However, many other past studies have described the association between the thyroid dysfunction and the depression (16, 20–22, 31, 41–44), with focuses on the “current thyroid dysfunction” as exposure, and most of them found significant positive associations, i.e. the subjects with thyroid dysfunction will have a higher risk of getting depression. The results of these studies together with ours indicate that the thyroid dysfunction may impact depression not only in a “short-term effect” but also in a “long-term effect” manner. Though there seems not many studies investigating the gender differences in the relationship between thyroid dysfunction and depression, one recently published paper (24) showed that the gender affects the association of thyroid-stimulating hormone levels with depression. The authors used data from the 2014 Korea National Health and Nutritional Examination Survey (KNHANES) as data source and found that the highest TSH level group was 1.92 times more likely to have depressive symptoms in males, but that number was 35% lower in females--the findings about the heterogeneity of the association between the thyroid dysfunction and the depression in different gender groups were generally in agreement with the findings from ours. They also claimed that this gender difference may be due to differences in thyroid response and autoimmunity (45) and the thyroid-stimulating hormone blunting and sex hormone levels (46), where this could also explain the findings in our study, even though their study focused on the current TSH levels as exposure.

### Strengths and Limitations

The presented study has the strength in analysis and interpretability. The use of NHANES data combining several collection years benefits the internal validity and the results from the design-adjusted analysis can be considered representative of the general US adults for the year 2013 to 2018. The screen instrument used to assess the depression (PHQ-9) is a validated instrument with adequate sensitivity and specificity (28). The variable ‘income’, which has the highest missing rate, was imputed as it is a variable that very likely to be “missing at random” (47), as people with lower income rate will be less likely to report (48). This imputation helps to reduce bias and the missing data analysis further aids to gain the estimation precision and the statistical power (47). The findings of this cross-sectional study can be considered relatively robust since the secondary analysis by multiple imputations and the sensitivity analysis by propensity score matching both showed similar and comparable results to the primary analysis.

However, the casual inference can not be stated due to the inherent limitation of the cross-sectional design and the available data does not allow for considerations of a temporal sequence. Another weakness is that most data in this study is based on self-reported measures which are subject to recall and report bias. For example, the smoking status is very likely to be under reported, especially for those who had thyroid dysfunction history—this will make the association between thyroid dysfunction history and depression overestimated for both male and female groups. Also, there are some known unmeasured confounding or risk factors of the outcome such as the genetic information that are found to be relevant to mood disorder (49), the recent major events such as the loss of a parent, etc. This information is not available, and it is hard to find good proxies for these variables in the NHANES data source—this unmeasured confounding issue may also make the association overestimated.

### Future direction

Future analysis should focus on investigating the relationship between the thyroid dysfunction history and the depression in other countries as the geographical (external) validation would strength the findings from this study. Also, unmeasured confounding especially the genetic information that are potentially relevant to the mood disorder should be assessed as it may affect the findings greatly. Further investigations of the association between the time-to-recovery of thyroid dysfunction and the depression could also be meaningful if the measures of time-to-recovery (i.e., when did the patients recovered) are available in other datasets.

### Implication

In summary, our study indicates that the thyroid dysfunction history can still impact the development of depression even though the subjects are recovered from the thyroid dysfunction. This impact has gender difference and is more severe for males than females. The higher risk of getting depression among males with thyroid dysfunction history are possibly due to the autoimmunity, the TSH blunting and sex hormone levels (46). This finding may help the health organizations with their policies/suggestions makings to prevent depression--more prevention suggestions and actions are needed for those who recovered from the thyroid dysfunction especially for the males.

## Data Availability

All data produced are available online at the National Health and Nutrition Examination Survey from 2013 to 2018.

https://wwwn.cdc.gov/nchs/nhanes/Default.aspx

## Notes

### Competing Interest Statement

The authors have declared no competing interest.

### Funding Statement

CIHR trainee award

### Author Declarations

The study used (or will use) ONLY openly available human data that were originally located at: National Health and Nutrition Examination Survey from 2013 to 2018.

